# AI-Based Clinical Decision Support Systems for Secondary Caries on Bitewings: A Multi-Algorithm Comparison

**DOI:** 10.64898/2026.04.17.26350883

**Authors:** E.T. Chaves, J.T. Domburg, V.H. Digmayer Romero, N. van Nistelrooij, S. Vinayahalingam, D. Sezen-Hulsmans, F.M. Mendes, M.C. Huysmans, M.S. Cenci, G.d.S. Lima

## Abstract

**Background:** Radiographic detection of caries lesions adjacent to restorations is challenging due to limitations of two-dimensional imaging and difficulties distinguishing true lesions from restorative or anatomical radiolucencies. Artificial intelligence (AI)-based clinical decision support systems (CDSSs) have been introduced to assist radiographic interpretation; however, different AI tools may yield variable diagnostic outputs, and their comparative performance remains unclear.

**Objective:** To compare the diagnostic performance of commercial and experimental AI algorithms for detecting secondary caries lesions on bitewings.

**Methods:** This cross-sectional diagnostic accuracy study included 200 anonymized bitewings comprising 885 restored tooth surfaces. A consensus group reference standard identified all surfaces with a caries lesion and classified each lesion by type (primary/secondary) and depth (enamel-only/dentin-involved). Five commercial (Second Opinion®, CranioCatch, Diagnocat, DIO Inteligência, and Align™ X-ray Insights) and three experimental (Mask R-CNN-based and Mask DINO-based) systems were tested. Diagnostic performance was expressed through sensitivity, specificity, and overall accuracy (95% CI). Comparisons used generalized estimating equations, adjusted for clustered data.

**Results:** Specificity was high across all systems (0.957–0.986), confirming accurate recognition of non-carious surfaces, whereas sensitivity was moderate (0.327–0.487), reflecting frequent missed detections of enamel and dentin lesions. Accuracy ranged from 0.882 to 0.917, with no significant differences among models (p ≥ 0.05). Confounding factors, such as radiographic overlapping, marginal restoration defects, and cervical artifacts, were the main sources of misclassification.

**Conclusions:** AI algorithms, regardless of architecture or commercial status, showed similar diagnostic capabilities and a conservative detection profile, favoring specificity over sensitivity. Improvements in dataset diversity, labeling precision, and explainability may further enhance reliability for secondary caries detection.

**Clinical Significance:** AI-based CDSSs assist clinicians by providing consistent detection. Their high specificity is particularly valuable in minimizing unnecessary invasive treatments (overtreatment), though they should be used as adjuncts rather than a replacement for expert judgment.

## 1 Introduction

Secondary caries, defined as lesions developing adjacent to restorations or sealants, remains one of the main causes of restoration failure and replacement [1]. Early detection of these lesions extends the longevity of restorations and supports minimally invasive treatment strategies [2]. Radiographic interpretation, particularly from bitewings, is routinely used to assist clinicians in identifying such lesions. However, diagnostic accuracy depends heavily on the operator’s expertise and may be compromised by radiographic confounding factors such as overlapping structures and restorative material artefacts [3].

Artificial intelligence (AI) has emerged as a promising tool to address these limitations by providing standardized analyses. Deep learning models, particularly convolutional neural networks (CNNs), have shown potential to enhance caries detection by delivering consistent, automated assessments [4,5]. Nevertheless, AI-based analyses remain sensitive to image quality and confounding variables, which can increase false-positive or false-negative results, emphasizing the importance of external validation before clinical adoption [6,7].

AI-based dental clinical decision support systems (CDSSs) mainly rely on two approaches: object detection and image segmentation. Object detection identifies and localizes specific regions of interest, whereas segmentation classifies each pixel of the image, enabling precise delineation of lesion boundaries [8,9]. Both approaches allow for distinction between carious and non-carious areas, and studies have reported satisfactory diagnostic accuracy for both primary and secondary caries, with sensitivities and specificities frequently exceeding 0.90 under experimental conditions [10].

Commercial AI-based CDSSs have demonstrated improvements in caries detection, particularly through increased sensitivity, although their impact on specificity remains inconsistent across studies [11]. This variability likely reflects differences in lesion characteristics, staging, and imaging conditions, as well as the inherent complexity of detecting caries adjacent to restorations. Evidence from both commercial and experimental models indicates that AI performance is highly context-dependent, with recurring trade-offs between improved detection and the risk of false-positive findings [5,11–16]. These patterns have been observed across different radiographic modalities, including bitewing and panoramic imaging, further supporting the influence of imaging conditions on diagnostic performance [15,16].

Despite the growing number of available algorithms, direct comparative studies among different models remain scarce. Differences in architecture, dataset composition, and calibration strategies may lead to distinct detection profiles with potential clinical implications. Some models may favor sensitivity, risking overtreatment through false positives, whereas others may emphasize specificity, increasing the likelihood of missed lesions (false negatives). From both clinical and regulatory perspectives, this variability highlights the need for transparent benchmarking, standardized datasets, and multicenter validation [17,18,19]. Additionally, for several AI systems, training data and code-level details are not publicly available, meaning that the mechanisms driving their diagnostic outputs still operate within a “black box” framework, potentially introducing algorithmic bias and reinforcing the importance of independent comparative evaluation.

Therefore, this study aimed to compare the diagnostic performance of commercial and experimental AI-based CDSSs for detecting secondary caries on bitewing radiographs using a shared, independent dataset. We hypothesized that differences in training data and model architectures would lead to different diagnostic accuracy across algorithms.

## 2 Material and Methods

### 2.1 Ethical considerations

This study complied with the principles of the Declaration of Helsinki. The manuscript was reported according to the reporting guideline for diagnostic accuracy studies using artificial intelligence (STARD-AI) [19]. Ethical approval was obtained from the CMO Arnhem-Nijmegen Ethics Committee (No. CMO 2025-17935). The study protocol was preregistered on the Open Science Framework (OSF) before data collection and is publicly available at https://n9.cl/l0ppu. All bitewings were anonymized before analysis to ensure confidentiality. No identifiable patient data were used.

### 2.2 Study design

This is a prospective diagnostic accuracy study comparing the performance of commercial and experimental AI-based dental clinical decision support systems (CDSSs) for detecting secondary caries on bitewings.

### 2.3 Dataset and participants

A total of 200 anonymized bitewing radiographs were selected from a database of clinical images obtained from dental clinics in Germany, the Netherlands, and Slovakia. Images were eligible if they had adequate image quality and depicted at least one restored permanent premolar or molar; only these teeth were included in the analysis. Radiographs were excluded if they exhibited motion artifacts, severe distortions, orthodontic or surgical appliances, duplicate images, or image height below 400 pixels. Each bitewing contributed multiple restored tooth surfaces, which were considered the unit of analysis. The dataset reflected typical clinical variation in restorative materials, including composite resin, amalgam, ceramic, and glass-ionomer restorations.

### 2.4 Reference standard

The reference standard was established through a structured expert-annotation process involving researchers and clinicians with expertise in cariology and radiographic image interpretation. Initially, a PhD candidate (ETC) and a cariology specialist (MSC) independently examined all bitewing radiographs under standardized viewing conditions, classifying each visible surface (mesial, occlusal, and distal) as non-carious, primary caries, or secondary caries. Each identified lesion was also staged by depth, using the enamel-dentin junction (EDJ) as the threshold, and categorized as enamel-only or dentin-involved. Subsequently, a review board composed of two additional PhD researchers (VHDR, DSH) and two experienced clinicians (GSL, MSC) re-evaluated all annotations. Any discrepancies between initial examiners were discussed and resolved by consensus. The final dataset was used as the reference standard for evaluating the diagnostic performance of all AI algorithms.

The choice of an expert consensus-based reference standard was based on the diagnostic nature of the bitewing images provided in this dataset and the need to reflect clinical decision-making. Following current paradigms in diagnostic research [20], expert consensus provides a pragmatic and reliable proxy for evaluating AI performance in real-world radiographic screening scenarios, despite the inherent risk of underestimating or overestimating lesion depth relative to invasive methods.

### 2.5 Index tests

Eight AI-based CDSSs for radiographic detection were grouped into commercial and experimental categories. The commercial systems comprised Second Opinion® (Pearl Inc., Los Angeles, USA), CranioCatch (CranioCatch, Eskisehir, Turkey), Diagnocat (Diagnocat, San Francisco, USA), DIO Inteligência (DIO Inteligência Odontológica, São Paulo, Brazil), and Align™ X-ray Insights (Align Technology Inc., San Jose, USA). These systems share the goal of assisting clinicians, though they differ in their interfaces and output formats. Some platforms provide region-level detections (e.g., bounding boxes or contours), while others add pixel-wise overlays around suspicious areas. All commercial CDSSs operate as proprietary, cloud-based solutions; their training datasets, preprocessing pipelines, calibration strategies, and post-processing rules are not publicly disclosed. For the present head-to-head comparison, each commercial system was used in its standard clinical configuration and evaluated on the same test set, without any additional tuning.

The experimental CDSSs (Exp_1, Exp_2, Exp_3) were developed by the Radboudumc Dental AI Hub (Nijmegen, The Netherlands). Exp_1 and Exp_2 implement Mask R-CNN with a Swin Transformer backbone [21,22], targeting instance-level segmentation of caries and restorations, whereas Exp_3 uses a hierarchical Mask DINO framework for multi-class segmentation with explicit lesion staging [15]. These academic models, including their training/validation regimes and architectural choices, have been previously described in the literature. None of the experimental models was trained on images from the present independent test set.

### 2.6 Data processing

Each bitewing was individually uploaded to the AI platforms. The outputs were independently annotated by two examiners (ETC, HD), who were blinded to the reference standard decisions. Both annotators followed the same decision framework used to establish the ground truth, recording for each tooth surface the presence or absence of a lesion, its type (primary or secondary), and, when available, its depth (enamel-only or dentin involvement). For CDSSs that did not explicitly differentiate between primary and secondary caries, the presence of a restoration adjacent to the detected lesion served as the threshold for classifying it as a secondary lesion. Similarly, for models that did not provide lesion staging, the enamel-dentin junction (EDJ) was used as the reference to categorize the lesion as either enamel-only or dentin-involved.

After the first round of independent annotations, the two examiners exchanged their datasets for cross-verification, comparing and discussing potential discrepancies. Any disagreements were resolved by consensus, with an experienced cariology expert (MSC) involved when necessary. All final diagnostic outputs, along with the validated annotated dataset, were securely archived and are publicly accessible through the OSF repository.

### 2.7 Statistical analysis

All analyses were conducted using Stata/SE 15.0 (StataCorp, College Station, USA) and RStudio (R Foundation, Vienna, Austria). Inter-examiner reliability was determined by Cohen’s κ, calculated separately for lesion type (non-carious, primary, secondary) and lesion depth (enamel, dentin). Agreement between each AI-based CDSS and the reference standard was quantified by percentage concordance and Cohen’s κ with respective 95% confidence intervals (95% CI). The frequency of indeterminate results (e.g., inconclusive classifications or no prediction outputs) was compared between each AI method and the reference standard using multilevel logistic regression, considering (tooth and bitewing as hierarchical levels).

Diagnostic performance was assessed through sensitivity, specificity, and overall accuracy (95% CI), adjusted for clustering of teeth within bitewings. Pairwise comparisons across CDSSs were tested using generalized estimating equations (GEE) with a logit link and exchangeable correlation structure, and estimated marginal means and p-values were computed using the emmeans package in R (v. 4.5.1). A secondary multilevel logistic regression model assessed the effects of tooth type (premolar vs. molar), arch (upper vs. lower), side (right vs. left), and surface (proximal vs. occlusal) on diagnostic outcomes, expressed as odds ratios (95% CI). A significance level of p < 0.05 was adopted for all analyses.

## 3. Results

A total of 200 bitewings were analyzed, comprising 885 restored tooth surfaces. Among these, the reference standard identified 115 (13%) secondary caries lesions, 12 (1.4%) of which were limited to enamel, and 103 (12%) reached the dentin, while 747 (84%) surfaces were considered as non-carious and 23 (2.6%) were indeterminate. The inter-examiner agreement for the reference standard was substantial (κ = 0.69 for lesion type and κ = 0.65 for lesion depth). All eight AI algorithms were tested under identical conditions for head-to-head comparison.

The frequency distributions (Figure 1; Supp Table 1) showed that all CDSSs most frequently classified restored surfaces as non-carious, consistent with the reference annotations. For enamel lesions, detections were limited: Second Opinion® and CranioCatch each identified 3/12, DiagnoCat 4/12, and both DIO Inteligência and Align™ X-ray 5/12. Most errors were downgrades to non-carious rather than overstaging to dentin, indicating a clear underestimation bias, with missed early lesions (false negatives) rather than overestimating healthy surfaces (false positives). For dentin lesions, performance varied more widely. Second Opinion® achieved the highest recall (51/103), followed by DiagnoCat (41/103) and DIO Inteligência (34/103), while CranioCatch (28/103) and Align™ X-ray (27/103) detected the fewest. In all cases, misclassifications were predominantly downward (dentin classified as non-carious), reinforcing the general trend of underdetection with few overstaging errors.

**Figure 1.**
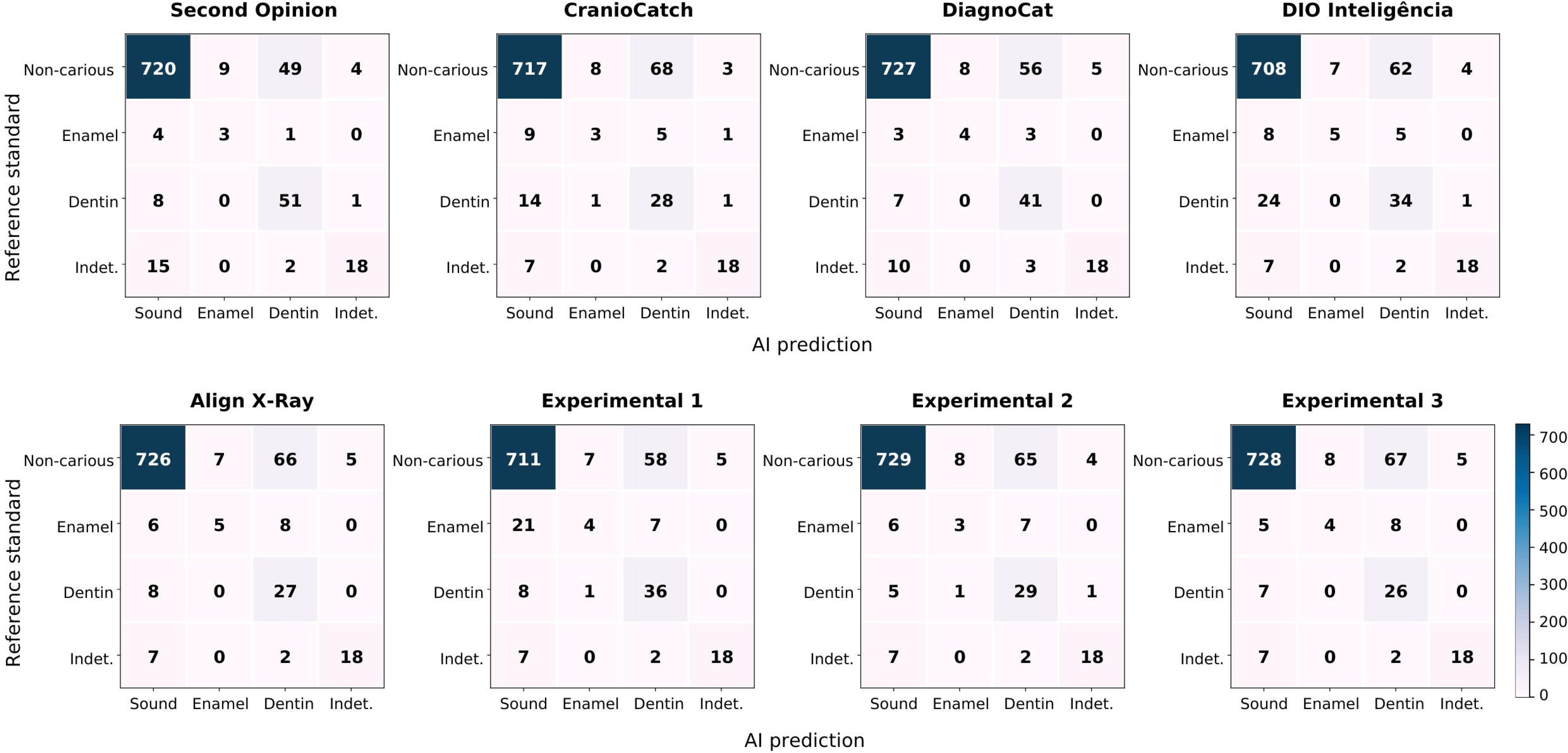
Confusion matrices illustrating the distribution of predictions from dental clinical decision support systems for secondary caries detection.

The experimental models showed a similar detection profile. For enamel lesions, Exp_1 detected 4/12, Exp_2 3/12, and Exp_3 4/12, with nearly all remaining cases labeled as non-carious (false negatives), confirming the challenge of recognizing early, low-contrast lesions even under standardized conditions. For dentin lesions, Exp_1 detected 36/103, Exp_2 29/103, and Exp_3 26/103, with most undetected lesions downgraded to non-carious rather than enamel.

Agreement with the reference standard was moderate across models, with concordance ranging from 86.4% (DIO Inteligência) to 89.5% (Second Opinion®) and κ values between 0.45 and 0.57 (95% CI) (Table 2). Among commercial systems, Second Opinion® achieved the highest κ (0.57) and accuracy (91.7%), while CranioCatch (κ = 0.43; accuracy = 88.4%) and DIO Inteligência (κ = 0.45; accuracy = 88.2%) showed slightly lower agreement. The experimental models performed comparably (κ = 0.45–0.47; accuracy = 89–90%), indicating consistent detection behavior across architectures.

**Table 1.** Agreement (% concordance) and Cohen’s kappa value (95% confidence intervals – 95%CI) between the CDSS classifications and the reference standard.

| CDSS | Reference Standard |  | Indeterminate results (%) |
| --- | --- | --- | --- |
|  | Concordance (%) | Kappa value (95%CI) |  |
| **Second Opinion®** | 89.5 | 0.571<br>(0.520 to 0.607) | 35 (4.0) |
| **CranioCatch** | 86.6 | 0.425<br>(0.388 to 0.487) | 31 (3.5) |
| **DiagnoCat** | 89.3 | 0.540<br>(0.460 to 0.551) | 27 (3.1) |
| **DIO Inteligência** | 86.4 | 0.450 | 27 (3.1) |
|  |  | (0.398 to 0.508) |  |
| **Align™ X-ray** | 87.7 | 0.459<br>(0.418 to 0.495) | 27 (3.1) |
| **Exp_1 [21]** | 86.9 | 0.471<br>(0.386 to 0.555) | 27 (3.1) |
| **Exp_2 [22]** | 88.0 | 0.469<br>(0.446 to 0.492) | 27 (3.1) |
| **Exp_3 [15]** | 87.7 | 0.450<br>(0.412 to 0.490) | 27 (3.1) |
| Artificial intelligence (AI); Dental Clinical Decision Support System (CDSS) |  |  |  |

**Table 2.** Accuracy parameters of CDSSs for detecting lesions adjacent to restorations.

| **CDSS** | **Specificity**<br>**(95%CI)** | **Sensitivity**<br>**(95%CI)** | **Accuracy**<br>**(95%CI)** |
| --- | --- | --- | --- |
| **Second Opinion®** | 0.984<br>(0.971 to 0.991) | 0.487<br>(0.387 to 0.587) | 0.917<br>(0.893 to 0.936) |
| **CranioCatch** | 0.969<br>(0.952 to 0.980) | 0.327<br>(0.250 to 0.416) | 0.884<br>(0.857 to 0.906) |
| **DiagnoCat** | 0.986<br>(0.973 to 0.993) | 0.429<br>(0.336 to 0.527) | 0.913<br>(0.888 to 0.932) |
| **DIO Inteligência** | 0.957<br>(0.934 to 0.972) | 0.389<br>(0.302 to 0.485) | 0.882<br>(0.852 to 0.906) |
| **Align™ X-ray** | 0.981<br>(0.969 to 0.988) | 0.354<br>(0.274 to 0.443) | 0.898<br>(0.874 to 0.918) |
| **Exp_1 [21]** | 0.961 | 0.425 | 0.890 |
|  | (0.943 to 0.973) | (0.337 to 0.518) | (0.868 to 0.909) |
| **Exp_2 [22]** | 0.985<br>(0.972 to 0.992) | 0.354<br>(0.266 to 0.454) | 0.902<br>(0.876 to 0.922) |
| **Exp_3 [15]** | 0.984<br>(0.972 to 0.991) | 0.336<br>(0.255 to 0.429) | 0.898<br>(0.875 to 0.917) |

95%CI = 95% confidence interval adjusted by the cluster.
No statistically significant difference among the methods for the same parameters ( $p \geq 0.05$ ) was observed using generalized estimating equations (GEE).
Artificial intelligence (AI); Dental Clinical Decision Support System (CDSS).

**Table 3.** Multilevel logistic regression analyses investigating accuracy parameters of CDSSs for detecting lesions adjacent to restorations.

| Independent variable | Specificity | Sensitivity | Accuracy |
| --- | --- | --- | --- |
|  | Odds Ratio (95% Confidence Interval) |  |  |
| **Molar vs. premolar** | 1.03<br>(0.68 to 1.55) | 1.19<br>(0.80 to 1.78) | 0.93<br>(0.76 to 1.13) |
| **Lower arch vs. upper arch** | 0.84<br>(0.57 to 1.25) | 1.06<br>(0.74 to 1.52) | 0.96<br>(0.80 to 1.14) |
| **Left vs. right side** | 0.811<br>(0.42 to 1.58) | 1.41<br>(0.87 to 2.31) | 1.08<br>(0.72 to 1.63) |
| **Proximal vs. occlusal surface** | 4.43 *<br>(2.26 to 8.68) | 0.30 *<br>(0.20 to 0.45) | 0.21 *<br>(0.16 to 0.26) |

\* Association statistically significant ( $p < 0.05$ ).

All systems displayed high specificity (0.96–0.99), reliably identifying non-carious surfaces (true negatives), but lower sensitivity (0.33–0.49), reflecting frequent missed detections (false negatives). No statistically significant differences were observed among algorithms (p ≥ 0.05), suggesting similar diagnostic performance under identical testing conditions. Indeterminate classifications were rare (3.1–4.0%), close to the reference standard (2.6%), and did not significantly affect agreement (p ≥ 0.05).

Multilevel logistic regression (Table 4) showed no significant effects of tooth type (premolar vs. molar), arch (upper vs. lower), or side (right vs. left) on diagnostic outcomes (p ≥ 0.05). The distribution of surfaces was predominantly proximal (83%), reflecting the typical caries detection process through bitewings. However, surface type significantly influenced results: proximal surfaces presented higher specificity (odds ratio = 4.43; 95% CI: 2.26–8.68) but lower sensitivity (odds ratio = 0.30; 95% CI: 0.20–0.45) and accuracy (odds ratio = 0.21; 95% CI: 0.16–0.26) compared to occlusal surfaces. This indicates that while non-carious proximal surfaces were reliably identified (true negatives), enamel and dentin lesions in these regions were more frequently missed (false negatives), particularly when overlapping restorations or anatomical structures were present. This trend was consistent across all models, underscoring the persistent challenge of proximal secondary caries detection in bitewings, even with advanced AI assistance. These findings should be interpreted with caution, as surface distribution and radiographic detectability may have influenced the observed differences between proximal and occlusal surfaces.

Figure 2 illustrates the outputs of all AI systems on the same bitewing, selected for its multiple secondary caries lesions, identified by the reference standard. Despite analyzing the same image, the algorithms produced heterogeneous outputs, with some detecting only a few of the lesions while others produced detections into non-carious areas.

**Figure 2.**
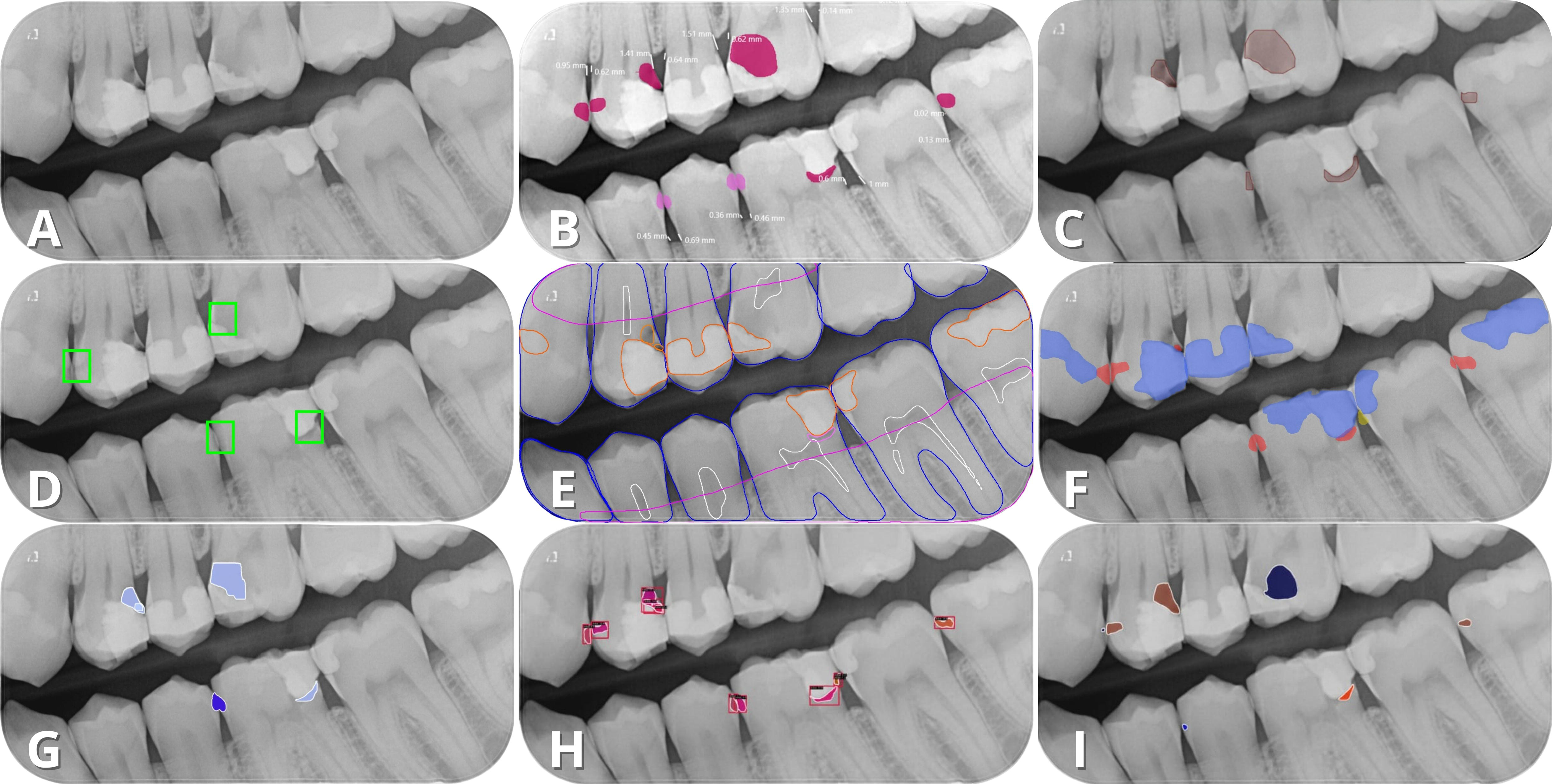
Panels A–I illustrate the outputs of each system on the same bitewing: A = original image; B = Second Opinion®; C = CranioCatch; D = DiagnoCat; E = Align™ X-ray Insights; F = DIO Inteligência; and G–I = experimental models (Exp_1-Exp_3).

Across all systems, the models favored specificity (true negatives) over sensitivity (true positives), reflecting a conservative diagnostic profile. Both commercial and experimental algorithms consistently underestimated lesion presence, mainly missing true lesions rather than overcalling non-carious areas. Although false-positive predictions were less frequent, they were clinically relevant, as many originated from radiographic confounders such as restoration defects, image artifacts, and overlapping structures. Figure 3 highlights examples of false-positive and false-negative detections caused by these confounding factors.

**Figure 3.**
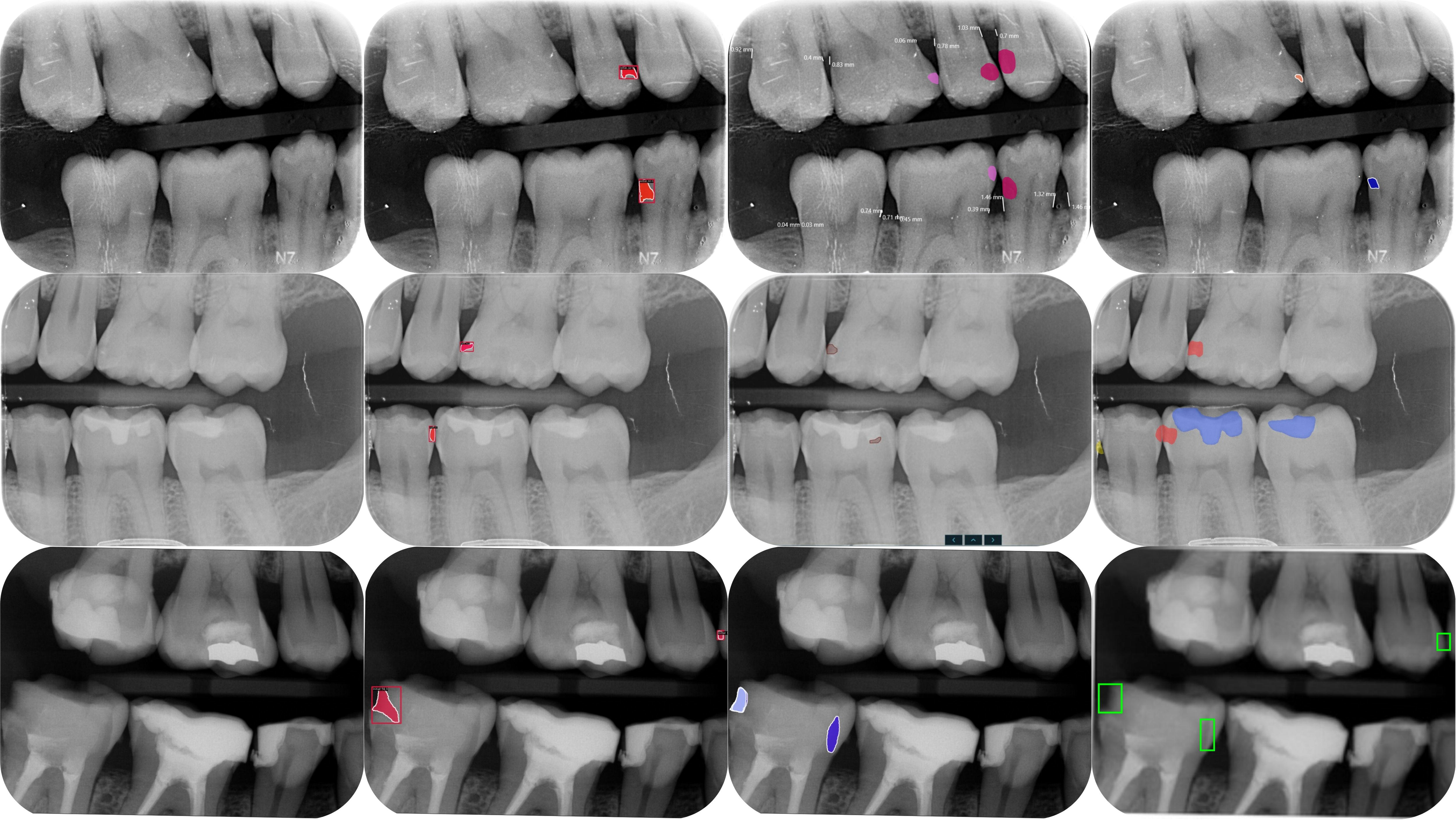
Representation of confounding factors. First row: radiolucent artifact at the distal mandibular second premolar. Second row: overlapping structures at the distal premolars (maxillary and mandibular). Third row: marginal defect at the distal mandibular second molar, all misclassified as lesions by multiple CDSSs.

## Discussion

This study investigated the hypothesis that differences in model architecture and training data would lead to distinct detection performances among different AI-based dental clinical decision support systems (CDSSs). Although some variability was observed, no statistically significant differences were found, thus refuting our working hypothesis. This outcome suggests that, once a certain level of technical maturity is reached, the performance of these CDSSs depends less on network architecture and more on dataset quality, labeling strategy, and clinical complexity.

Across all CDSSs, specificity remained consistently high (95.7–98.6%), confirming reliable identification of non-carious surfaces, whereas sensitivity was lower (32.7–48.7%). Commercial and experimental models performed within similar ranges (accuracy ≈ 88–92%), yet both exhibited a detection pattern characterized by high specificity and lower sensitivity. This pattern likely reflects the inherent difficulty of detecting secondary caries on bitewings, particularly in the presence of radiographic confounders and subtle lesion presentation. In a clinical workflow, these CDSSs may act as adjunctive tools to support decision-making and help prevent unnecessary restorative interventions, a key principle in minimally invasive dentistry. However, their limited sensitivity precludes their use as standalone triage systems, as a negative AI result cannot reliably exclude disease. Instead, these algorithms should be considered supportive tools to assist practitioners in identifying potentially overlooked lesions during chairside radiographic assessment, with final diagnostic decisions remaining dependent on the integration of clinical history and visual examination [2,3,6].

Balancing sensitivity and specificity remains a critical challenge in caries diagnostics. Improving sensitivity enhances the recognition of subtle or incipient lesions, while also increasing the likelihood of false positives, potentially leading to overtreatment and restorative cycles [20]. Previous studies have shown similar trade-offs between traditional and adjunctive diagnostic methods, in which combined visual and radiographic approaches improved lesion detection at the cost of reduced specificity and superfluous interventions [23,24]. Systematic reviews reinforce this pattern, emphasizing that radiography is more sensible for advanced lesions, offering limited accuracy for early-stage disease [25]. At the same time, validated visual methods maintain balanced performance and support minimally invasive care [26]. In this context, the diagnostic behavior observed across AI algorithms mirrors long-standing clinical trends: reliable recognition of advanced disease with limited sensitivity for early detection.

When compared to previous AI studies, our results align with evidence [27,28]. These investigations also show better performance of the models for primary caries compared to secondary caries. CNNs and object-detection frameworks often achieve high accuracy in detecting primary lesions, occasionally outperforming less experienced clinicians [7,13]. Meta-analyses have confirmed strong pooled sensitivity and specificity for primary caries detection [7], yet fewer studies have validated these systems for secondary lesions. In those that did, such as models based on Mask R-CNN or YOLO, reported sensitivities varied widely depending on the dataset and annotation protocol [14,15,16,21]. This variability highlights that dataset composition and image characteristics may have a greater impact on diagnostic performance than algorithmic architecture itself, a finding consistent with our results.

The difficulty in detecting secondary caries lesions in bitewings lies in the overlapping structures, restorative marginal defects, adhesive layers, and anatomical structures. These factors obscure lesion margins and produce image patterns similar to caries radiolucencies, leading to underdetection or misclassification. Our analysis confirmed that most misclassifications were downward shifts (true lesions classified as non-carious) rather than overcalls, reinforcing that confounding radiographic features remain the dominant barrier to accurate detection of secondary caries, even with the diagnostic outputs of the AI-based CDSSs.

Another key finding concerns transparency and accessibility. Many commercial CDSSs remain (partially) closed platforms; data sources, labeling protocols, and post-market performance indicators are not publicly available. This opacity limits reproducibility and regulatory evaluation, reinforcing the need for standardized benchmarking and open datasets. In contrast, most experimental models, often developed within academic frameworks, achieved comparable accuracy using transparent architectures and shareable data [13,15,21,22]. This suggests that diagnostic accuracy in secondary caries may depend more on domain-specific data quality and the relevance of the training set to the test environment than on the sheer volume of ‘big data’ available to commercial platforms. Experimental CDSSs, often trained on smaller yet highly curated datasets, demonstrated that localized calibration can match the performance of proprietary global algorithms. This reinforces the hypothesis that technological maturity has plateaued, and further gains in accuracy will likely require higher-quality reference standards rather than more complex architectures. This observation supports broader efforts toward open science and reproducibility in dental AI research.

Several methodological and algorithmic limitations should be acknowledged. First, the sample was drawn from a convenience dataset, selected to reflect realistic clinical variability rather than the population-level distribution. Second, the reliance on expert visual-radiographic consensus for both model training and as a reference standard may introduce a circularity bias. Since the AI models were developed to replicate human interpretation rather than histological truth, the observed performance likely reflects the models’ ability to mimic current clinical subjectivity and its inherent diagnostic limitations. This suggests that the ‘performance ceiling’ encountered across systems may be a byproduct of labeling strategies that propagate human observational gaps into algorithmic outputs. Third, differences in lesion prevalence and detectability may have influenced accuracy estimates, particularly for enamel-only lesions, which are difficult to visualize in bitewings. Similarly, secondary caries on occlusal surfaces may be obscured by structural superimposition. As a result, only more advanced lesions, typically involving dentin, were more likely to be identified, which may have influenced the observed diagnostic performance. At the algorithmic level, none of the models provided explainability metrics such as saliency maps or heatmaps, which could clarify their decision-making process. This “black-box” behavior limits interpretability and may perpetuate hidden biases, particularly when image artifacts correlate with class labels. Finally, regional restrictions on software availability prevented inclusion of some commercial platforms, illustrating ongoing inequities in AI deployment and evaluation.

Future research should move beyond retrospective comparisons toward prospective, clinically integrated trials that assess diagnostic thresholds, the impact on decision-making, and patient outcomes. Developing shared, annotated datasets specifically for secondary caries, and incorporating explainability frameworks to visualize AI reasoning, would greatly increase transparency and accelerate regulatory validation. Moreover, collaborative benchmarking between academic groups and commercial developers could establish common standards for performance reporting, error analysis, and continuous post-market evaluation.

## Conclusion

The tested artificial intelligence (AI)-based dental clinical decision support systems (CDSSs) showed comparable diagnostic performance for detecting secondary caries in bitewings, with high specificity and moderate sensitivity. Differences in architecture did not lead to significant performance gaps, suggesting that factors beyond network design, such as dataset characteristics, may influence diagnostic outcomes. Overall, the systems demonstrated consistent yet conservative diagnostic behavior, prioritizing the avoidance of false positives over the detection of early secondary caries lesions.

### Declaration of generative AI

During the preparation of this report, the authors used text generative AI tools to assist in improving language clarity and readability. All outputs were reviewed, edited, and validated by the authors, who take full responsibility for the final content of the manuscript.

## Data Availability

All data supporting the findings of this study are available at the Open Science Framework (OSF) repository: https://n9.cl/l0ppu

## Contributorship

**Conceptualization:** E.T.C., S.V., M.S.C., M.C.H., G.S.L.; **Methodology:** E.T.C., H.D., M.S.C., F.M.M., M.C.H., G.S.L.; **Formal analysis:** E.T.C., H.D., M.S.C., F.M.M., M.C.H.; **Investigation:** E.T.C., H.D., V.H.D.R., N.v.N., S.V., D.S.H.; **Data curation:** E.T.C., H.D., V.H.D.R., N.v.N., S.V., D.S.H.; **Writing – original draft:** E.T.C., H.D., M.S.C.; **Writing – review & editing:** E.T.C., H.D., G.S.L., F.M.M., M.S.C., M.C.H.; **Supervision:** G.S.L., F.M.M., M.S.C., M.C.H.

---

**Supp Table 1.**
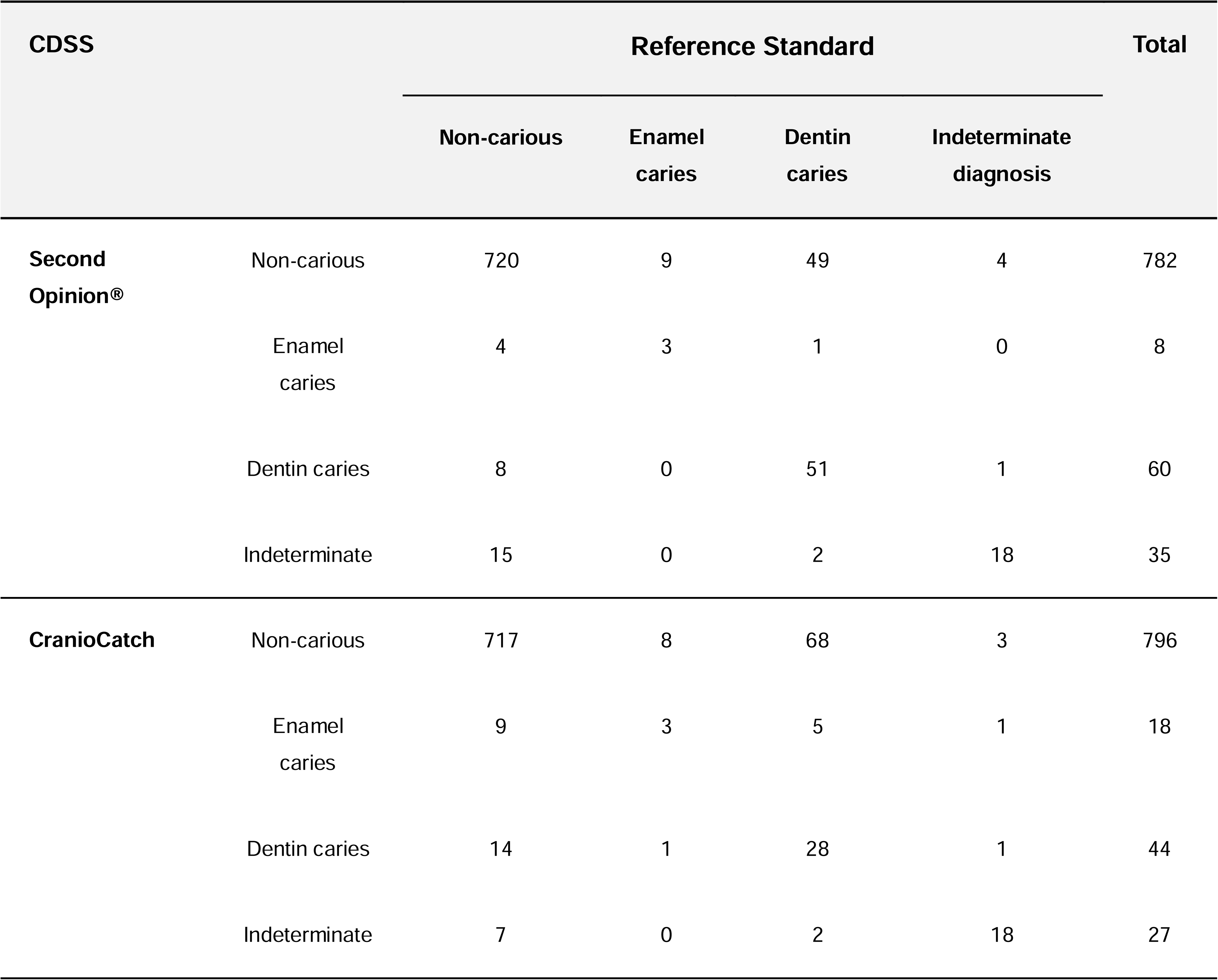
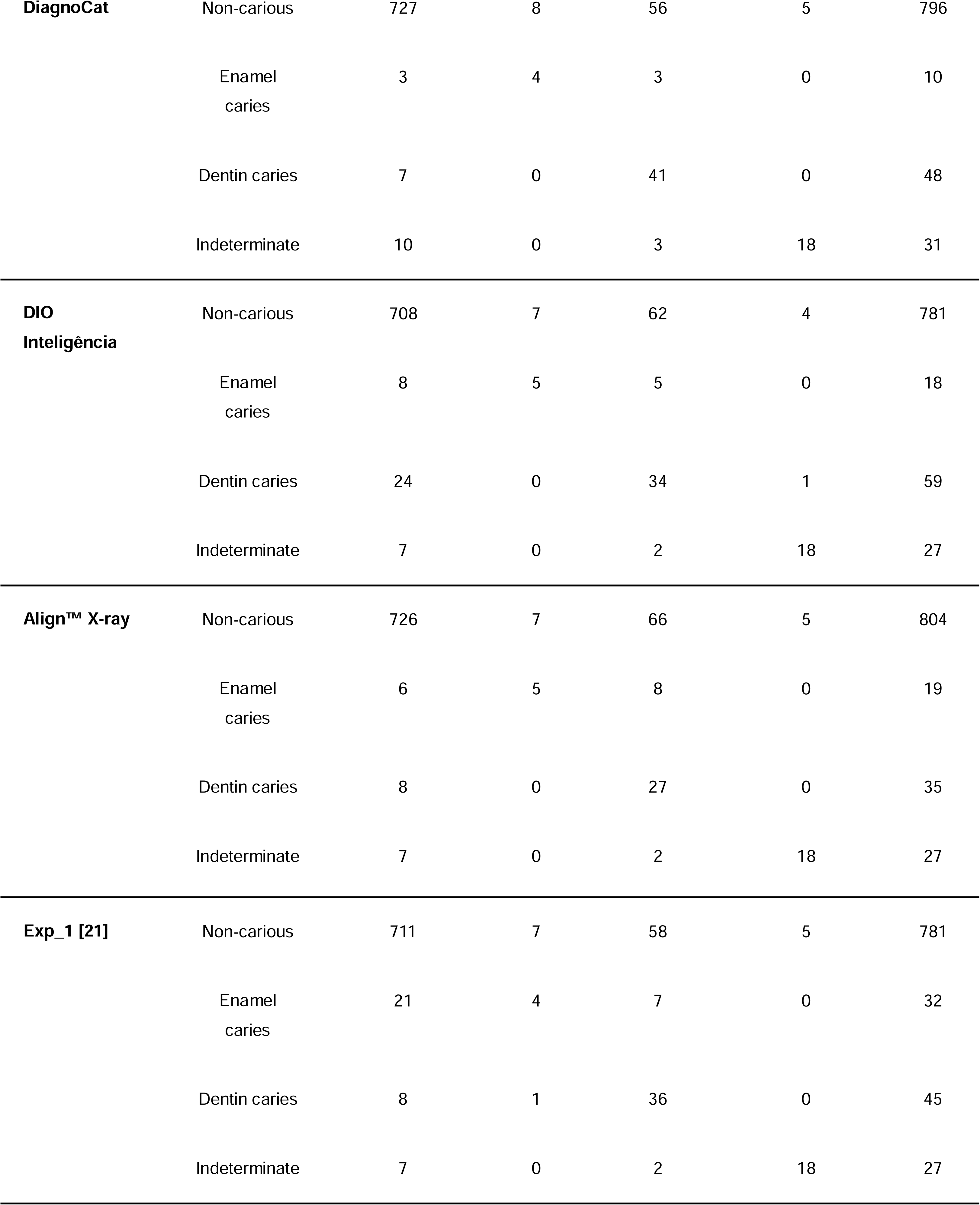
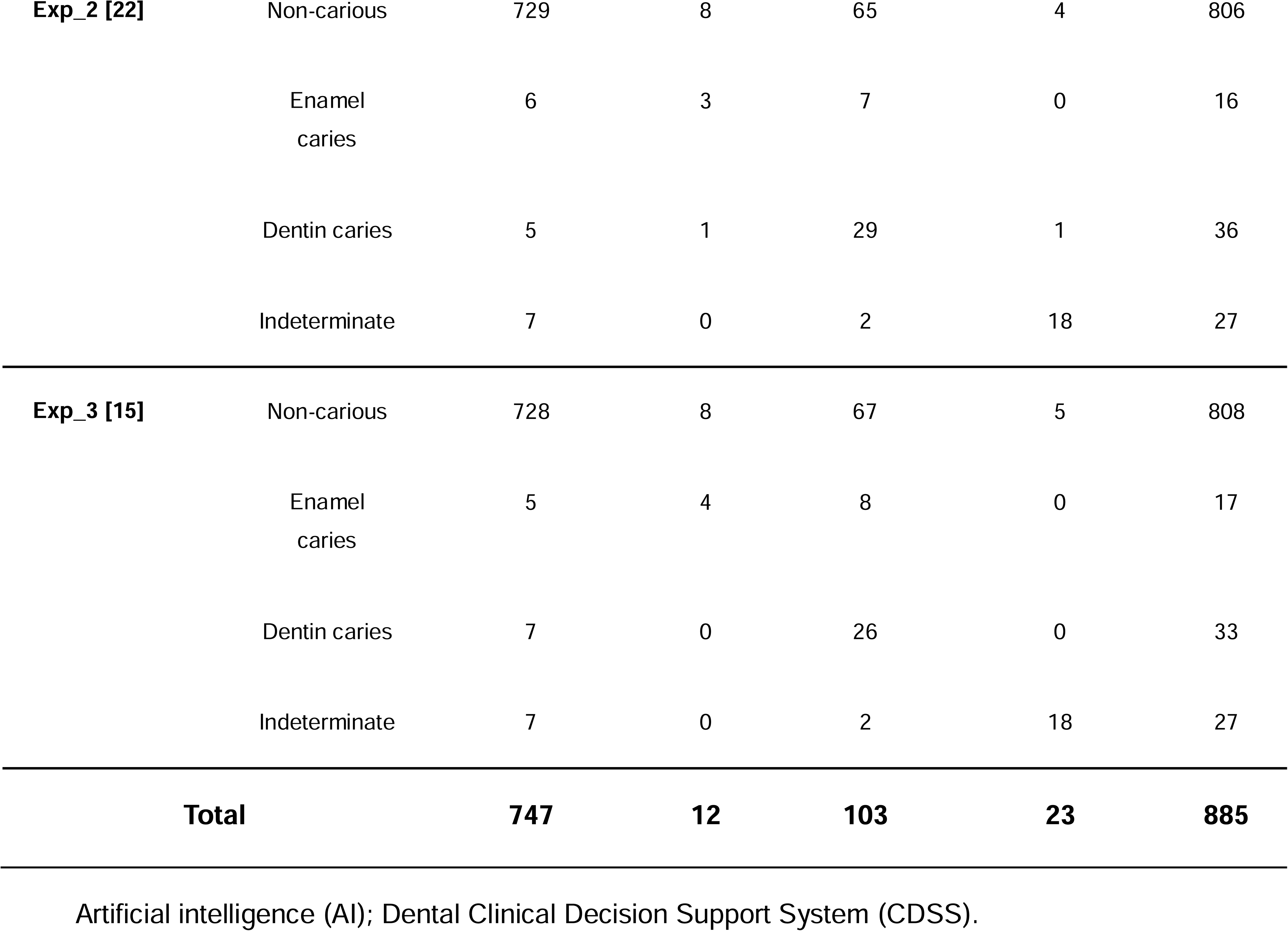
Contingency tables for the staging obtained by the CDSSs compared to the reference standard.

| CDSS |  | Reference Standard |  |  |  | Total |
| --- | --- | --- | --- | --- | --- | --- |
|  |  | Non-carious | Enamel caries | Dentin caries | Indeterminate diagnosis |  |
| **Second Opinion®** | Non-carious | 720 | 9 | 49 | 4 | 782 |
|  | Enamel caries | 4 | 3 | 1 | 0 | 8 |
|  | Dentin caries | 8 | 0 | 51 | 1 | 60 |
|  | Indeterminate | 15 | 0 | 2 | 18 | 35 |
| **CranioCatch** | Non-carious | 717 | 8 | 68 | 3 | 796 |
|  | Enamel caries | 9 | 3 | 5 | 1 | 18 |
|  | Dentin caries | 14 | 1 | 28 | 1 | 44 |
|  | Indeterminate | 7 | 0 | 2 | 18 | 27 |
| **DiagnoCat** | Non-carious | 727 | 8 | 56 | 5 | 796 |
|  | Enamel caries | 3 | 4 | 3 | 0 | 10 |
|  | Dentin caries | 7 | 0 | 41 | 0 | 48 |
|  | Indeterminate | 10 | 0 | 3 | 18 | 31 |
| **DIO**<br>**Inteligência** | Non-carious | 708 | 7 | 62 | 4 | 781 |
|  | Enamel caries | 8 | 5 | 5 | 0 | 18 |
|  | Dentin caries | 24 | 0 | 34 | 1 | 59 |
|  | Indeterminate | 7 | 0 | 2 | 18 | 27 |
| **Align™ X-ray** | Non-carious | 726 | 7 | 66 | 5 | 804 |
|  | Enamel caries | 6 | 5 | 8 | 0 | 19 |
|  | Dentin caries | 8 | 0 | 27 | 0 | 35 |
|  | Indeterminate | 7 | 0 | 2 | 18 | 27 |
| **Exp_1 [21]** | Non-carious | 711 | 7 | 58 | 5 | 781 |
|  | Enamel caries | 21 | 4 | 7 | 0 | 32 |
|  | Dentin caries | 8 | 1 | 36 | 0 | 45 |
|  | Indeterminate | 7 | 0 | 2 | 18 | 27 |
| **Exp_2 [22]** | Non-carious | 729 | 8 | 65 | 4 | 806 |
|  | Enamel<br>caries | 6 | 3 | 7 | 0 | 16 |
|  | Dentin caries | 5 | 1 | 29 | 1 | 36 |
|  | Indeterminate | 7 | 0 | 2 | 18 | 27 |
| **Exp_3 [15]** | Non-carious | 728 | 8 | 67 | 5 | 808 |
|  | Enamel<br>caries | 5 | 4 | 8 | 0 | 17 |
|  | Dentin caries | 7 | 0 | 26 | 0 | 33 |
|  | Indeterminate | 7 | 0 | 2 | 18 | 27 |
| **Total** |  | **747** | **12** | **103** | **23** | **885** |

Artificial intelligence (AI); Dental Clinical Decision Support System (CDSS).

